# Incident psychopharmacological treatment and psychiatric hospital contact in individuals with newly developed type 2 diabetes

**DOI:** 10.1101/2020.07.21.20158733

**Authors:** Christopher Rohde, Norbert Schmitz, Reimar W. Thomsen, Søren D. Østergaard

## Abstract

**Objective:** To investigate the association between newly developed type 2 diabetes (T2D) and incident psychopharmacological treatment and psychiatric hospital contact.

**Methods:** We identified all individuals from the Central- and Northern Denmark Regions with newly developed T2D (defined by the first HbA_1c_ measurement >6.5%) from 2000-2016 and up to five age and sex matched individuals without T2D (controls). Those having received psychopharmacological treatment or having had a psychiatric hospital contact in the five years prior to the onset of T2D were excluded. For this cohort, we first assessed the incidence of psychopharmacological treatment and psychiatric hospital contact among individuals with T2D and controls, respectively. Secondly, we compared the incidence of psychopharmacological/psychiatric hospital contact among individuals with T2D to propensity score matched controls. Finally, we assessed which baseline (T2D onset) characteristics that were associated with subsequent psychopharmacological treatment and psychiatric hospital contact.

**Results:** We identified 56,640 individuals with newly developed T2D and 315,694 controls. A total of 8.3% of the individuals with T2D initiated psychopharmacological treatment within the 2 years following onset compared to 4.6% among the age and sex matched controls. Individuals with T2D were at increased risk of initiating psychopharmacological treatment compared to the propensity score matched controls (HR=1.51, 95% CI=1.43-1.59), whereas their risk of psychiatric hospital contact was not increased to the same extent (HR=1.14, 95% CI=0.98-1.32). Older age, somatic comorbidity, and being divorced/widowed was associated with both psychopharmacological treatment and psychiatric hospital contact following T2D.

**Conclusion:** Individuals with T2D are at elevated risk of requiring psychopharmacological treatment.

**Significant outcomes:**

- 8.3% of the individuals with T2D initiated psychopharmacological treatment within the 2 years following onset compared to 4.6% among the age and sex matched controls
- Individuals with newly developed T2D were at increased risk of initiating psychopharmacological treatment and of having psychiatric hospital contact compared to propensity score matched controls.
- Risk factors for psychopharmacological treatment/psychiatric hospital contact following development of T2D include older age, somatic comorbidity and being divorced or widowed.

**Limitations:**

- Identification of T2D (a HbA1c level >6.5%) itself might lead to the identification of mental illness and thereby psychopharmacological treatment initiation/psychiatric hospital contact.
- A proportion of the individuals with T2D will likely have initiated treatment with an antidepressant due to neuropathic pain developed as a complication to T2D

## Introduction

Type 2 diabetes (T2D) is associated with a range of severe complications including micro- and macrovascular disease, neuropathy, cancer, dementia, and infections (Nwaneri et al., 2013, Harding et al., 2019a). In addition, there is mounting evidence suggesting that psychiatric comorbidity is also highly prevalent in T2D (Semenkovich et al., 2015, Harding et al., 2019b). Specifically, prior studies have shown that individuals with T2D are twice as likely to exhibit symptoms of depression, anxiety, insomnia, and distress compared to the general population (Semenkovich et al., 2015, Collins et al., 2009, Khuwaja et al., 2010, Smith et al., 2013, Tan et al., 2018, Hackett and Steptoe, 2017, Ducat et al., 2014) - with up to one out of five developing major depression (Semenkovich et al., 2015). Accordingly, prior studies have established that individuals with T2D are more likely to use antidepressants than individuals from the general population (Manderbacka et al., 2011); with the prevalence of antidepressant use in T2D ranging between 7-10% (Mast et al., 2017, Ivanova et al., 2010, Manderbacka et al., 2011). In addition, the prevalence of anxiolytic/hypnotic has been estimated at 7% (Mast et al., 2017). It is unclear whether this high prevalence of psychiatric symptoms and psychopharmacological treatment stems from psychological distress and biological alternations caused by T2D or is due to existing psychiatric illness (Kim et al., 2016, Khuwaja et al., 2010). Another possibility is that T2D primarily is an innocent marker of poor socioeconomic status, somatic comorbidity, obesity, and unhealthy lifestyle, which all are risk factors for/associated with mental illness (Kolb and Martin, 2017).

Prior studies that have investigated psychopharmacological treatment and need for psychiatric hospital care in individuals with T2D are mainly limited by three things. First, studies aiming at studying these aspects in individuals with newly developed T2D have often not had the exact date of T2D diagnosis at their disposal, and therefore focused on selected populations using T2D treatment initiation or hospitalization as a proxy for the onset of T2D (Mast et al., 2017, Cleal et al., 2018). Second, prior studies have generally not been able to take preexisting mental illness or psychopharmacological treatment into account (Mast et al., 2017, Perez et al., 2017, Cleal et al., 2018, Kivimäki et al., 2010), i.e. whether the need for treatment arose before or after the development of T2D. Third, adjustment for potential confounders of the association between T2D and psychopharmacological treatment requires access to information that has not been available in many prior studies (Mast et al., 2017, Perez et al., 2017, Cleal et al., 2018, Kivimäki et al., 2010).

As the limitations outlined above may have resulted in inaccurate estimates of incident psychiatric morbidity in individuals with newly developed T2D, we conducted a register-based study aimed at overcoming these limitations. This was done by i) defining onset of T2D as the first measured HbA1c level >6.5% (48mmol/mol), ii) excluding individuals having received psychopharmacological treatment or having had a psychiatric hospital contact in the five years preceding the onset of T2D, and iii) conducting a propensity score matched analysis of the association between newly developed T2D and incident psychopharmacological treatment and psychiatric hospital contact.

## Material and Methods

### Design and setting

We conducted a register-based study based on a dataset containing information on all individuals residing in the Northern Denmark and Central Denmark Regions (approximately 1.9 million inhabitants). The registers providing data for the study are described below.

### Registers

The Danish Civil Registration System (DCRS) was established in 1968 and contains the unique personal registration numbers, which are assigned to all individuals living in Denmark (at birth or when becoming a legal resident), enabling linkage of data at the individual from all public registers (Pedersen, 2011). In the current study, we linked data from the following registers: 1) The Information System (LABKA) database, which contains laboratory results from all general practitioners and hospitals in the Central and Northern Denmark Regions since the 1990s with full completeness since the early 2000s (Grann et al., 2011). 2) The Danish National Patient Register (DNPatR), which contains discharge diagnoses from all admissions to Danish non-psychiatric hospitals since 1977 and from emergency and outpatient hospital settings since 1995 (Lynge et al., 2011). 3) The Danish Psychiatric Central Research Register (DPCRR), which contains discharge diagnoses from all psychiatric hospital admissions since 1969 and from emergency and outpatient hospital settings since 1995 (Mors et al., 2011). 4) The Danish National Prescription Register (DNPreR), which contains date on prescriptions redeemed at all Danish pharmacies since 1995 (Kildemoes et al., 2011).

### Study population

LABKA was used to identify all individuals with onset of T2D in the period from January 1, 2000 to October 31, 2015 (defined as the first date with a blood sample with a HbA1c level >6.5% (48mmol/mol)).

Individuals that prior to this date had redeemed a prescription for a glucose lowering drug (see definition in Supplementary Table 1), had a register-based diagnosis of T2D (see definition in Supplementary Table 1), or were under 30 years old at the time of diagnosis were excluded to ensure that the exact date for T2D onset was known and to minimize the presence of individuals with type 1 diabetes in the dataset (Mor et al., 2016). Subsequently, to enable investigation of incident psychopharmacological/psychiatric hospital treatment, individuals with psychopharmacological/psychiatric hospital treatment (see definition in Supplementary Table 1) in the five years preceding the onset of T2D were excluded. The remaining individuals with T2D were matched on age sex and date (onset of T2D) with up to 5 individuals without T2D (controls) and without psychopharmacological/psychiatric hospital treatment in the five years preceding the matched date. Individuals included as matched controls could be included in another matched pair (as an individual with T2D) if he/she developed T2D after the matched date.

### Propensity score matching

In an attempt to minimize the degree of confounding in the estimation of the association between T2D and psychopharmacological treatment/psychiatric hospital contact, we carried out a propensity score matched analysis. Specifically, individuals with T2D from the cohort described above were matched 1:1 to one individual from the age and sex matched control group on covariates (see section below) associated with T2D and psychiatric illness/psychopharmacological treatment. All covariates that were expected to be associated with exposure and outcome (all of those described below) were used for the propensity score matching. The first step in the matching procedure was to calculate the predicted probability (the propensity score) of each individual in the cohort having T2D on the basis of his or her covariate profile, using logistic regression. Subsequently, the propensity scores were trimmed asymmetrically using the 95^th^ percentile of the propensity scores for individuals without T2D and the 5^th^ percentile for individuals with T2D (Stürmer et al., 2010). We then matched each individual with T2D to the individual without T2D having the propensity score closest to that of the former using calipers of width equal to 0.2 of the standard deviation (Austin, 2011). The matching was checked with a kernel density plot and with the mean standardized difference (see Figure 2).

**Table 1:**
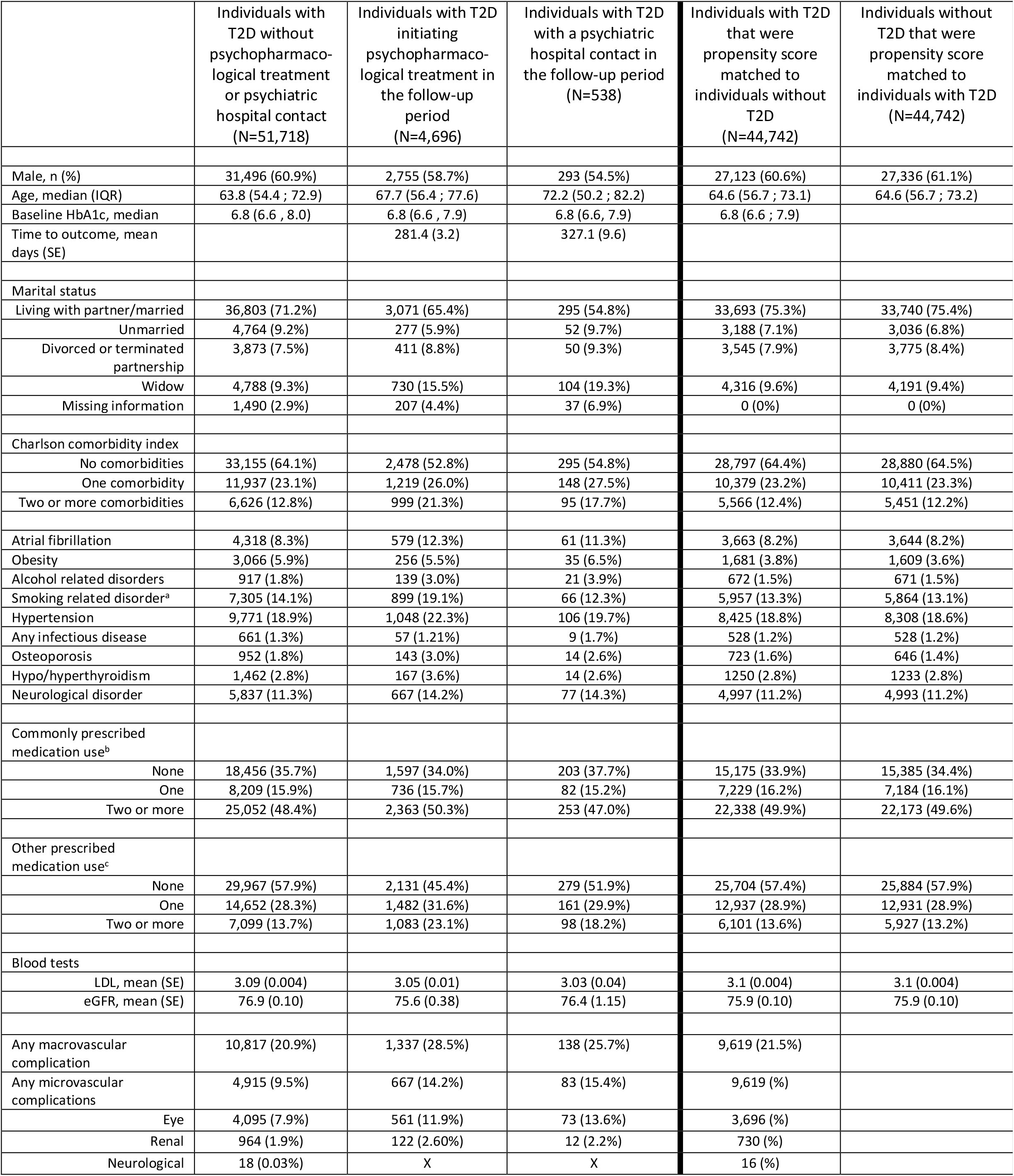
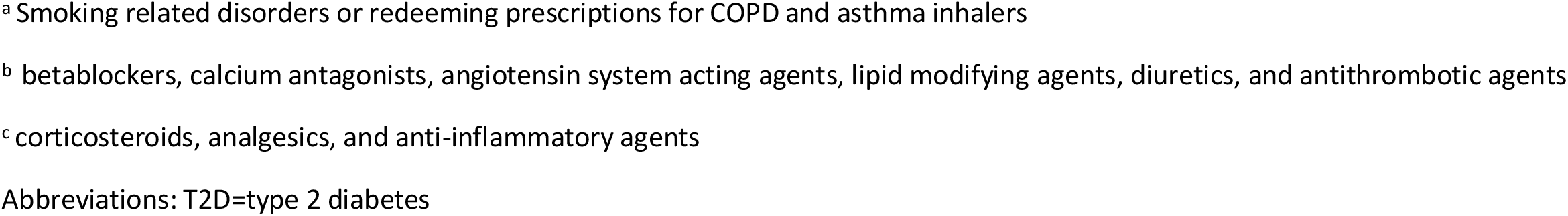
Baseline characteristics of individuals with T2D (stratified on psychopharmacological treatment and psychiatric hospital contact status) and propensity score matched controls without T2D.

|  | Individuals with T2D without psychopharmacological treatment or psychiatric hospital contact (N=51,718) | Individuals with T2D initiating psychopharmacological treatment in the follow-up period (N=4,696) | Individuals with T2D with a psychiatric hospital contact in the follow-up period (N=538) | Individuals with T2D that were propensity score matched to individuals without T2D (N=44,742) | Individuals without T2D that were propensity score matched to individuals with T2D (N=44,742) |
| --- | --- | --- | --- | --- | --- |
| Male, n (%) | 31,496 (60.9%) | 2,755 (58.7%) | 293 (54.5%) | 27,123 (60.6%) | 27,336 (61.1%) |
| Age, median (IQR) | 63.8 (54.4 ; 72.9) | 67.7 (56.4 ; 77.6) | 72.2 (50.2 ; 82.2) | 64.6 (56.7 ; 73.1) | 64.6 (56.7 ; 73.2) |
| Baseline HbA1c, median | 6.8 (6.6 , 8.0) | 6.8 (6.6 , 7.9) | 6.8 (6.6, 7.9) | 6.8 (6.6 ; 7.9) |  |
| Time to outcome, mean days (SE) |  | 281.4 (3.2) | 327.1 (9.6) |  |  |
| Marital status |  |  |  |  |  |
| Living with partner/married | 36,803 (71.2%) | 3,071 (65.4%) | 295 (54.8%) | 33,693 (75.3%) | 33,740 (75.4%) |
| Unmarried | 4,764 (9.2%) | 277 (5.9%) | 52 (9.7%) | 3,188 (7.1%) | 3,036 (6.8%) |
| Divorced or terminated partnership | 3,873 (7.5%) | 411 (8.8%) | 50 (9.3%) | 3,545 (7.9%) | 3,775 (8.4%) |
| Widow | 4,788 (9.3%) | 730 (15.5%) | 104 (19.3%) | 4,316 (9.6%) | 4,191 (9.4%) |
| Missing information | 1,490 (2.9%) | 207 (4.4%) | 37 (6.9%) | 0 (0%) | 0 (0%) |
| Charlson comorbidity index |  |  |  |  |  |
| No comorbidities | 33,155 (64.1%) | 2,478 (52.8%) | 295 (54.8%) | 28,797 (64.4%) | 28,880 (64.5%) |
| One comorbidity | 11,937 (23.1%) | 1,219 (26.0%) | 148 (27.5%) | 10,379 (23.2%) | 10,411 (23.3%) |
| Two or more comorbidities | 6,626 (12.8%) | 999 (21.3%) | 95 (17.7%) | 5,566 (12.4%) | 5,451 (12.2%) |
| Atrial fibrillation | 4,318 (8.3%) | 579 (12.3%) | 61 (11.3%) | 3,663 (8.2%) | 3,644 (8.2%) |
| Obesity | 3,066 (5.9%) | 256 (5.5%) | 35 (6.5%) | 1,681 (3.8%) | 1,609 (3.6%) |
| Alcohol related disorders | 917 (1.8%) | 139 (3.0%) | 21 (3.9%) | 672 (1.5%) | 671 (1.5%) |
| Smoking related disorder<sup>a</sup> | 7,305 (14.1%) | 899 (19.1%) | 66 (12.3%) | 5,957 (13.3%) | 5,864 (13.1%) |
| Hypertension | 9,771 (18.9%) | 1,048 (22.3%) | 106 (19.7%) | 8,425 (18.8%) | 8,308 (18.6%) |
| Any infectious disease | 661 (1.3%) | 57 (1.21%) | 9 (1.7%) | 528 (1.2%) | 528 (1.2%) |
| Osteoporosis | 952 (1.8%) | 143 (3.0%) | 14 (2.6%) | 723 (1.6%) | 646 (1.4%) |
| Hypo/hyperthyroidism | 1,462 (2.8%) | 167 (3.6%) | 14 (2.6%) | 1250 (2.8%) | 1233 (2.8%) |
| Neurological disorder | 5,837 (11.3%) | 667 (14.2%) | 77 (14.3%) | 4,997 (11.2%) | 4,993 (11.2%) |
| Commonly prescribed medication use<sup>b</sup> |  |  |  |  |  |
| None | 18,456 (35.7%) | 1,597 (34.0%) | 203 (37.7%) | 15,175 (33.9%) | 15,385 (34.4%) |
| One | 8,209 (15.9%) | 736 (15.7%) | 82 (15.2%) | 7,229 (16.2%) | 7,184 (16.1%) |
| Two or more | 25,052 (48.4%) | 2,363 (50.3%) | 253 (47.0%) | 22,338 (49.9%) | 22,173 (49.6%) |
| Other prescribed medication use<sup>c</sup> |  |  |  |  |  |
| None | 29,967 (57.9%) | 2,131 (45.4%) | 279 (51.9%) | 25,704 (57.4%) | 25,884 (57.9%) |
| One | 14,652 (28.3%) | 1,482 (31.6%) | 161 (29.9%) | 12,937 (28.9%) | 12,931 (28.9%) |
| Two or more | 7,099 (13.7%) | 1,083 (23.1%) | 98 (18.2%) | 6,101 (13.6%) | 5,927 (13.2%) |
| Blood tests |  |  |  |  |  |
| LDL, mean (SE) | 3.09 (0.004) | 3.05 (0.01) | 3.03 (0.04) | 3.1 (0.004) | 3.1 (0.004) |
| eGFR, mean (SE) | 76.9 (0.10) | 75.6 (0.38) | 76.4 (1.15) | 75.9 (0.10) | 75.9 (0.10) |
| Any macrovascular complication | 10,817 (20.9%) | 1,337 (28.5%) | 138 (25.7%) | 9,619 (21.5%) |  |
| Any microvascular complications | 4,915 (9.5%) | 667 (14.2%) | 83 (15.4%) | 9,619 (%) |  |
| Eye | 4,095 (7.9%) | 561 (11.9%) | 73 (13.6%) | 3,696 (%) |  |
| Renal | 964 (1.9%) | 122 (2.60%) | 12 (2.2%) | 730 (%) |  |
| Neurological | 18 (0.03%) | X | X | 16 (%) |  |

<sup>a</sup> Smoking related disorders or redeeming prescriptions for COPD and asthma inhalers
<sup>b</sup> betablockers, calcium antagonists, angiotensin system acting agents, lipid modifying agents, diuretics, and antithrombotic agents
<sup>c</sup> corticosteroids, analgesics, and anti-inflammatory agents
Abbreviations: T2D=type 2 diabetes

**Figure 1:**
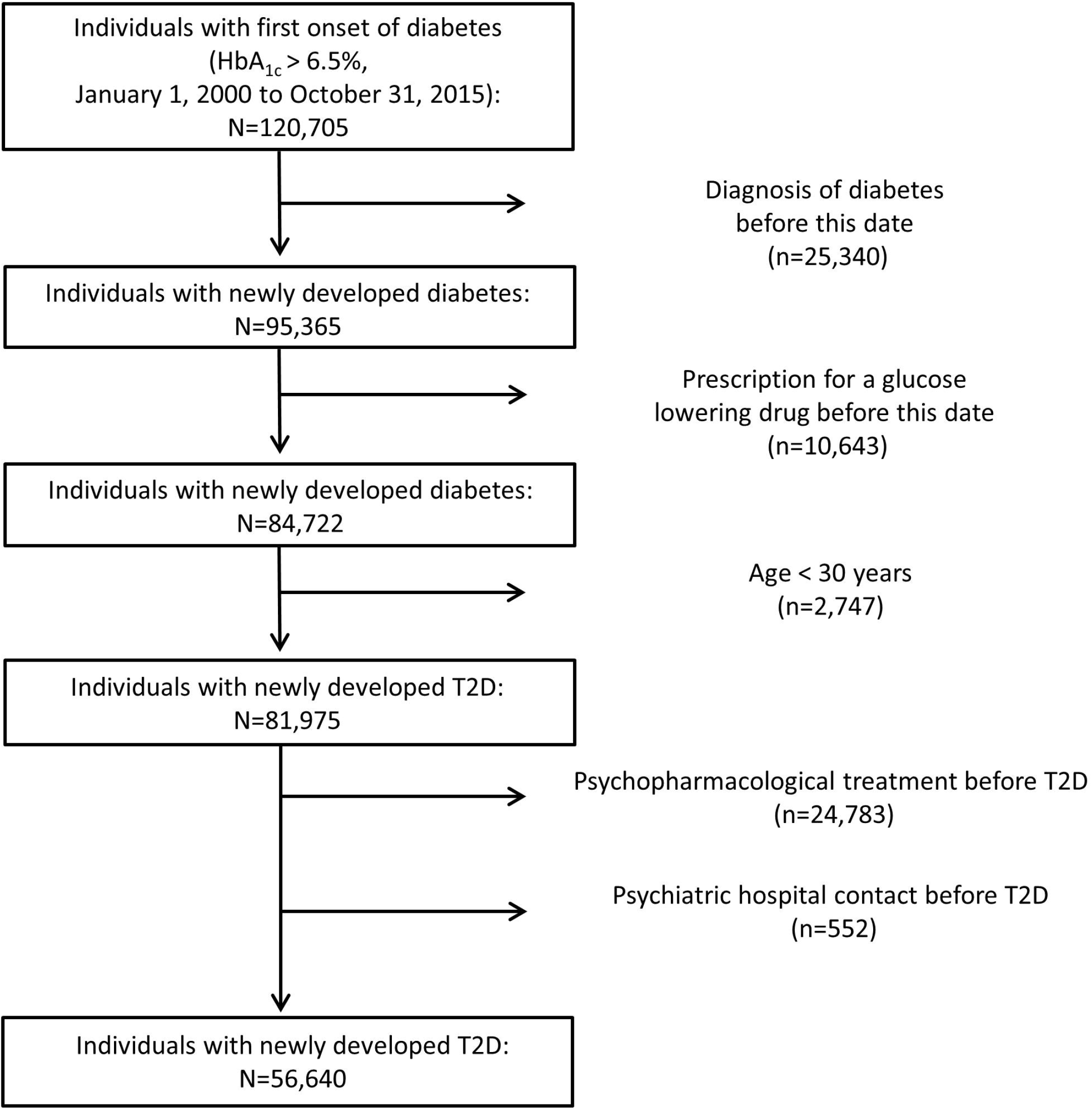
Selection of study population. Abbreviations: T2D=type 2 diabetes

**Figure 2:**
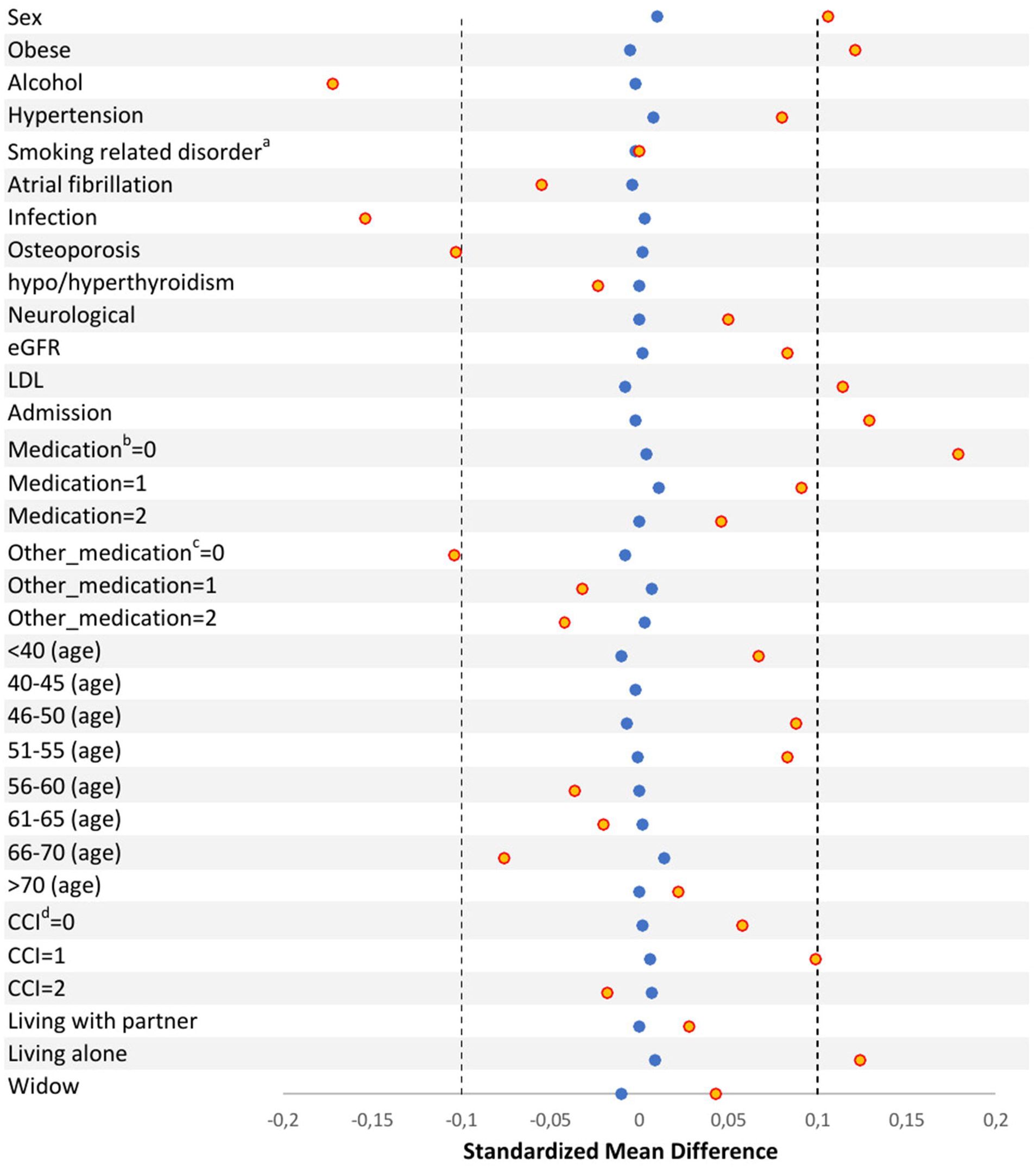
Mean standardized differences before (orange color) and after (blue color) propensity score matching. ^a^ Smoking related disorder or redemption of a prescription for a drug used in the treatment of chronic obstructive airway disease (COPD) or asthma. ^b^ betablockers, calcium antagonists, renin-angiotensin system acting agents, lipid modifying agents, diuretics, and antithrombotic agents ^c^ Including corticosteroids, analgesics, and anti-inflammatory agents ^c^ Charlson comorbidity index.

### Covariates

We obtained data regarding a number of covariates, which were used to calculate the propensity score described above and as potential predictors of incident psychopharmacological treatment and psychiatric hospital contact. Specifically, information on sex, age, and marital status at the onset of T2D/matched date were extracted from the DCRS. Information on baseline HbA_1c_, LDL, and eGFR levels were extracted from LABKA. Information on the 17 major disease groups included in the Charlson Comorbidity Index (excluding diabetes and diabetes with chronic complications) were obtained from the DNPatR and the cohort members were categorized based on the number of diseases registered at baseline (0, 1, and ≥2). In addition to the diseases included in the Charlson Comorbidity Index, information on atrial fibrillation, hypertension, infectious diseases, osteoporosis, alcohol related disorders, obesity, hyper/hypothyroidism, neurological disorders, and micro- and macrovascular complications were obtained from the DNPatR. As information on smoking behavior is not available in the Danish registers, we defined a “smoking related disorder” covariate based on data from the DNPatR (see definition in Supplementary Table 1) and the DNPreR (redemption of a prescription for a drug used in the treatment of obstructive airway disease). Furthermore, information on redeemed prescriptions for the most commonly used medications for somatic illnesses (betablockers, calcium antagonists, renin-angiotensin system acting agents, lipid modifying agents, diuretics, and antithrombotic agents) in the one year preceding the onset of T2D was extracted from the DNPreR. The same was the case for medications with potential relation to T2D and/or mental illness (corticosteroids, analgesics, and anti-inflammatory medication). The definition of all covariates is provided in Supplementary Table 1.

### Outcomes

The outcomes were incident psychopharmacological treatment and psychiatric hospital contact, respectively. Psychopharmacological treatment initiation was considered a proxy for mild/moderate mental illness, whereas psychiatric hospital contact was considered to be associated with more severe psychiatric illness. The outcomes were not mutually exclusive, i.e. an individual could have both.

Psychopharmacological treatment was identified using the DNPreR and separate analyses were conducted for antidepressants, antipsychotics, and anxiolytics, respectively (see definitions in Supplementary Table 1). Psychiatric hospital contact was identified using the DPCRR. In addition to considering psychiatric hospital contact in general (resulting in a diagnosis in the mental disorder chapter of the International Classification of Diseases, 10^th^ Revision (ICD-10)), we also conducted analyses on contact resulting in diagnoses of depression, anxiety disorder, psychotic disorder, and substance use disorder, specifically (see ICD-10 definitions in Supplementary Table 1).

## Statistical Analyses

### Incidence rates of psychopharmacological treatment and psychiatric hospital contact

The cohort members were followed from the onset of T2D or the matched date until death, emigration, first psychopharmacological medication prescription, first psychiatric hospital contact, or two years from onset/matched date, whichever came first. If the controls developed T2D (HbA_1c_ >6.5%) they were censored at this date. For this follow-up period, the incidence rates of psychopharmacological treatment and psychiatric hospital contact were reported for the T2D-and the age and sex matched controls, respectively.

### The association between T2D and psychopharmacological treatment and psychiatric hospital contact

For this analysis, the individuals with T2D and the propensity score matched controls individuals were followed from the onset of T2D or the matched date until death, emigration, first psychopharmacological medication prescription, first psychiatric hospital contact, or two years from onset/matched date, whichever came first. If the controls developed T2D (HbA_1c_ >6.5%) they were censored at this date. We then carried out a matched Cox regression analysis with psychopharmacological treatment and psychiatric hospital contact (see definitions under “outcomes“).

### Risk factors for psychopharmacological treatment and psychiatric hospital contact in T2D

Here, we assessed which baseline covariates (see earlier) that were associated with subsequent psychopharmacological treatment and psychiatric hospital contact among individuals with T2D using Cox-proportional hazards regression. The follow-up was identical to that for the analyses described above. The proportional hazard assumptions were tested by plotting the observed survival curves against the estimated survival curves.

## Sensitivity Analyses

We repeated the analyses of incident psychopharmacological treatment by redefining the outcome to require redemption of at least two prescriptions for psychopharmacological drugs within the two years following onset of T2D/matched data. This was chosen, as redemption of at least two prescriptions may be a more specific marker of mental illness as compared to redemption of only one prescription (McCrea et al., 2016, Liu et al., 2017).

## Results

### Study population

The identification of the study population is illustrated in the flowchart in Figure 1. In brief, we identified 120,705 individuals with incident diabetes (defined by the HbA_1c_). From these, 35,983 had a redeemed a prescription for a glucose lowering drug or were diagnosed with diabetes at a hospital before this date and among the remaining individuals, 2,747 were <30 years old, leaving 81,975 individuals with newly developed T2D. From these, 25,335 individuals had received psychopharmacological/psychiatric hospital treatment in the five years leading up to the date of the T2D diagnosis and were excluded from the dataset to allow for investigation of (5-year) incident psychopharmacological treatment and psychiatric hospital contact. These steps led to the final cohort of 56,640 individuals. A total of 315,694 age and sex matched individuals without diabetes and who had not received psychopharmacological treatment or had any psychiatric hospital contact in the five years prior to the matched date were identified from the general population (in Central- and Northern Denmark). From the 315,694 controls, we were able to propensity score match a total of, 44,742 individuals to cohort members with T2D. The characteristics of the cohort with newly developed T2D, the proportion of the cohort with T2D that was propensity score matched, as well as the propensity scored matched controls from the general population are shown in Table 1.

### Incidence rates of psychopharmacological treatment and psychiatric hospital contact

Among the individuals with newly developed T2D, 8.3% initiated psychopharmacological treatment during follow-up. 5.6%, 1.2%, and 2.9% initiated treatment with an antidepressant-, antipsychotic-, or an anxiolytic agent, respectively (not mutually exclusive outcomes). A total of 1.0% of the individuals with newly developed T2D had a psychiatric hospital contact in the follow-up period. Among the age and sex matched controls from the general population, 4.6% initiated psychopharmacological treatment during follow-up. 2.6%, 0.6%, and 2.0% initiated treatment with an antidepressant-, antipsychotic-, or an anxiolytic agent, respectively. A total of 0.7% of the controls had a psychiatric hospital contact in the follow-up period (Table 2).

**Table 2:** Incident psychopharmacological treatment and psychiatric hospital contact in the first two years after incident T2D or matched date (for the controls)

|  | Individuals with T2D<br>N=56,640 (%) |  | Individuals without T2D (controls)<br>N=315,694 (%) |  |
| --- | --- | --- | --- | --- |
|  | Number of<br>individuals (%) | Incidence rate pr<br>1000 years follow-up,<br>(95% CI) | Number of<br>individuals (%) | Incidence rate pr 1000<br>years follow-up,<br>(95% CI) |
| **Any**<br>**psychopharmacological**<br>**agent** | 4,696 (8.3%) | 45.65 (44.36-46.97) | 14,539 (4.6%) | 23.97 (23.58-24.36) |
| **Antidepressants** |  |  |  |  |
| Any antidepressant | 3,159 (5.6%) | 30.37 (29.33-31.44) | 8,407 (2.7%) | 13.74 (13.44-14.04) |
| SSRI | 1,698 (3.0%) | 16.10 (15.35-16.89) | 4,453 (1.4%) | 7.24 (7.03-7.45) |
| SNRI | 229 (0.4%) | 2.14 (1.88-2.43) | 430 (0.1%) | 0.69 (0.63-0.76) |
| TCA | 655 (1.2%) | 6.15 (5.70-6.64) | 1,533 (0.5%) | 2.48 (2.36-2.61) |
| Other | 795 (1.4%) | 7.47 (6.97-8.01) | 2,329 (0.7%) | 3.77 (3.62-3.93) |
| **Antipsychotics** |  |  |  |  |
| Any antipsychotic | 703 (1.2%) | 6.59 (6.12-7.10) | 1,769 (0.6%) | 2.86 (2.73-3.00) |
| Olanzapine | 78 (0.1%) | 0.73 (0.58-0.91) | 207 (0.1%) | 0.33 (0.29-0.38) |
| Quetiapine | 107 (0.2%) | 1.00 (0.83-1.21) | 249 (0.1%) | 0.40 (0.36-0.46) |
| Risperidone | 131 (0.2%) | 1.22 (1.03-1.45) | 380 (0.1%) | 0.61 (0.55-0.67) |
| Haloperidol | 237 (0.4%) | 2.21 (1.95-2.51) | 455 (0.1%) | 0.73 (0.67-0.81) |
| **Anxiolytics** |  |  |  |  |
| Any anxiolytic | 1,643 (2.9%) | 15.52 (14.79-16.29) | 6,400 (2.0%) | 10.42 (10.17-10.68) |
| Benzodiazepines | 1,530 (2.7%) | 14.44 (13.73-15.18) | 6,057 (1.9%) | 9.86 (9.61-10.11) |
| **Psychiatric hospital**<br>**contact:** |  |  |  |  |
| Any contact<sup>a</sup> | 538 (1.0%) | 5.05 (4.64-5.49) | 2,334 (0.7%) | 3.78 (3.63-3.94) |
| Depressive disorder | 142 (0.3%) | 1.33 (1.13-1.56) | 613 (0.2%) | 0.99 (0.91-1.07) |
| Anxiety disorder | 22 (0.0%) | 0.21 (0.14-0.31) | 96 (0.0%) | 0.16 (0.13-0.19) |
| Psychotic disorder | 37 (0.1%) | 0.35 (0.25-0.48) | 145 (0.1%) | 0.23 (0.20-0.27) |
| Substance use disorder | 16 (0.0%) | 0.15 (0.09-0.24) | 88 (0.0%) | 0.14 (0.12-0.18) |

<sup>a</sup> Admission, outpatient contact, psychiatric emergency room contact.
Abbreviations: T2D=type 2 diabetes, SSRI=selective serotonin reuptake inhibitor, SNRI=serotonin–norepinephrine reuptake inhibitor, TCA=tricyclic antidepressant

### The association between T2D and psychopharmacological treatment and psychiatric hospital contact

A total of 7.7% of the individuals with newly developed T2D initiated psychopharmacological treatment compared to 5.3% of the propensity score matched controls. The Cox regression analysis showed that Individuals with newly developed T2D were at substantially increased risk of initiating psychopharmacological treatment in general (HR=1.51, 95%CI=1.43-1.59), as well as antidepressant (HR=1.66, 95%CI=1.56-1.78), antipsychotic (HR=1.74, 95%CI=1.49-2.01), and anxiolytic (HR=1.30, 95%CI=1.20-1.42) treatment specifically, compared to the propensity score matched controls (Table 3). A total of 0.8% of the individuals with newly developed T2D had psychiatric hospital contact compared to 0.7% of the propensity score matched controls. The Cox regression indicated that individuals with newly developed T2D also had a slightly increased risk of having a psychiatric hospital contact (HR=1.14, 95%CI=0.98-1.32).

**Table 3:** Incident psychopharmacological treatment and psychiatric hospital contact within the first two years after incident T2D or matched date (for the propensity score matched controls)

|  | Individuals with T2D<br>(n=44,742) | Propensity score matched<br>controls without T2D<br>(n=44,742) | Hazard ratio (HR) |
| --- | --- | --- | --- |
| **Any psychopharmacological agent** | 3,452 (7.7%) | 2,354 (5.3%) | 1.51 (1.43-1.59) |
| **Antidepressants** |  |  |  |
| Any antidepressant | 2,283 (5.1%) | 1,418 (3.2%) | 1.66 (1.56-1.78) |
| SSRI | 1,192 (2.7%) | 710 (1.6%) | 1.73 (1.57-1.89) |
| SNRI | 149 (0.3%) | 80 (0.2%) | 1.91 (1.46-2.51) |
| TCA | 491 (1.1%) | 292 (0.7%) | 1.72 (1.49-1.99) |
| Other | 559 (1.3%) | 408 (0.9%) | 1.40 (1.24-1.60) |
| **Antipsychotics** |  |  |  |
| Any antipsychotic | 466 (1.0%) | 275 (0.6%) | 1.74 (1.49-2.01) |
| **Anxiolytics** |  |  |  |
| Any anxiolytic | 2,210 (4.9%) | 951 (2.1%) | 1.30 (1.20-1.42) |
| Benzodiazepines | 1,124 (2.5%) | 893 (2.0%) | 1.29 (1.18-1.41) |
| **Psychiatric hospital contact** |  |  |  |
| Any contact<sup>a</sup> | 354 (0.8%) | 319 (0.7%) | 1.14 (0.98-1.32) |

<sup>a</sup> Admission, outpatient contact, psychiatric emergency room contact.
Abbreviations: T2D=type 2 diabetes, SSRI=selective serotonin reuptake inhibitor, SNRI=serotonin–norepinephrine reuptake inhibitor, TCA=tricyclic antidepressant

### Risk factors for psychopharmacological treatment and psychiatric hospital contact in T2D

The following characteristics were clear and statistically significant risk factors for psychopharmacological treatment following onset of T2D: high age (71-80 years versus <=50 years; HR=1.17, 95%CI=1.05-1.31 and >80 years versus <=50 years; HR=1.46, 95%CI=1.28-1.66), Charlson Comorbidity Index score of 1 (HR=1.22, 95%CI=1.13-1.32), Charlson Comorbidity Index score of 2 (HR=1.69, 95%CI=1.52-1.87), macrovascular complications (HR=1.16, 95%CI=1.07-1.26), being divorced (HR=1.22, 95%CI=1.09-1.35), being widowed (HR=1.35, 95%CI=1.23-1.49), having a smoking related disorders (HR=1.18, 95%CI=1.09-1.28), and having an alcohol related diagnosis (HR=1.66, 95%CI=1.37-2.00). Never being married (HR=0.71, 95%CI=0.62-0.81), age 51-60 versus <=50 years (HR=0.86, 95%CI=0.78-0.95), age 61-70 versus <=50 years (HR=0.89, 95%CI=0.81-0.98) and use of other commonly prescribed medications (using two medications; HR=0.76, 95%CI=0.70-0.82) was associated with reduced likelihood of psychopharmacological treatment (see Table 4).

**Table 4:**
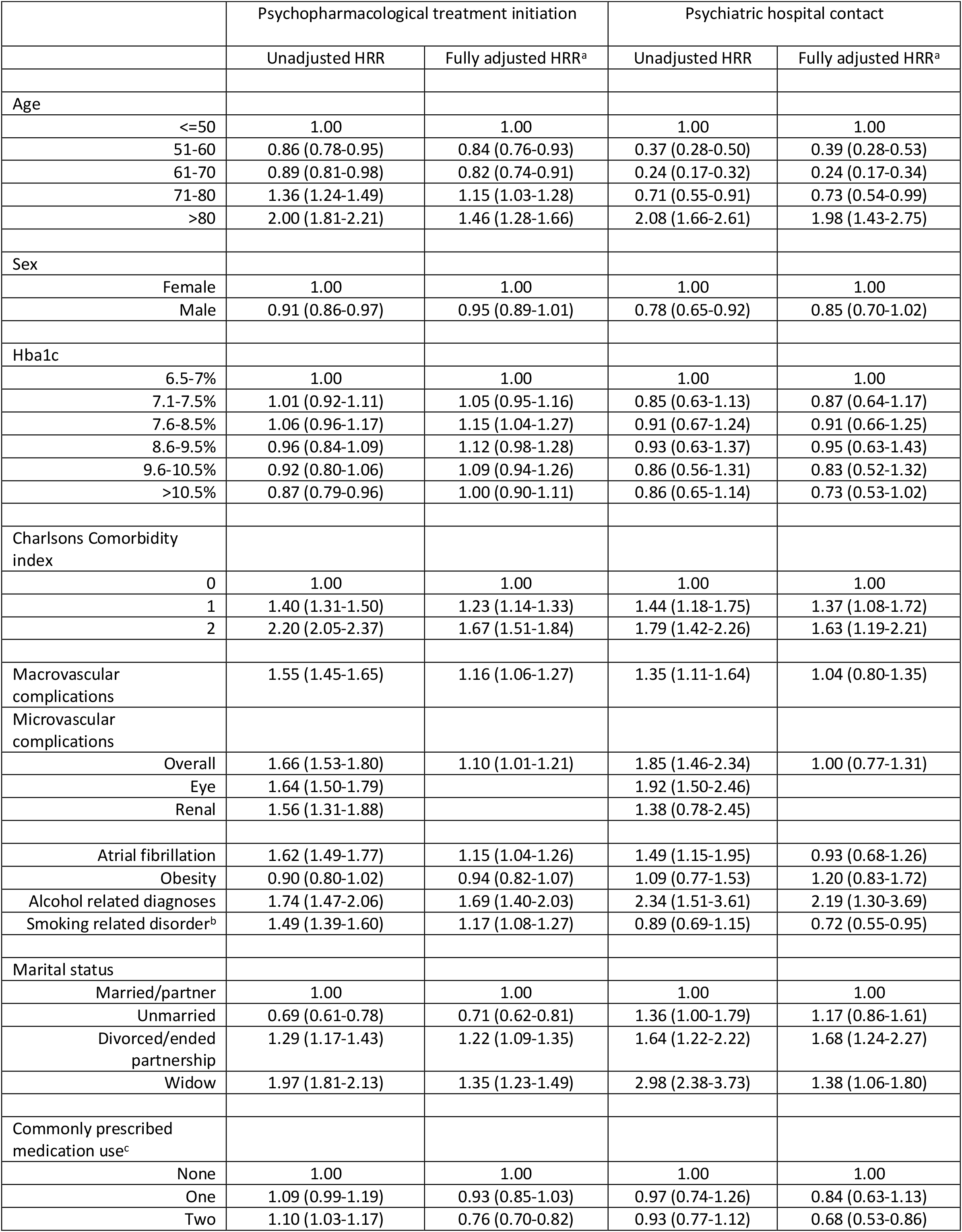
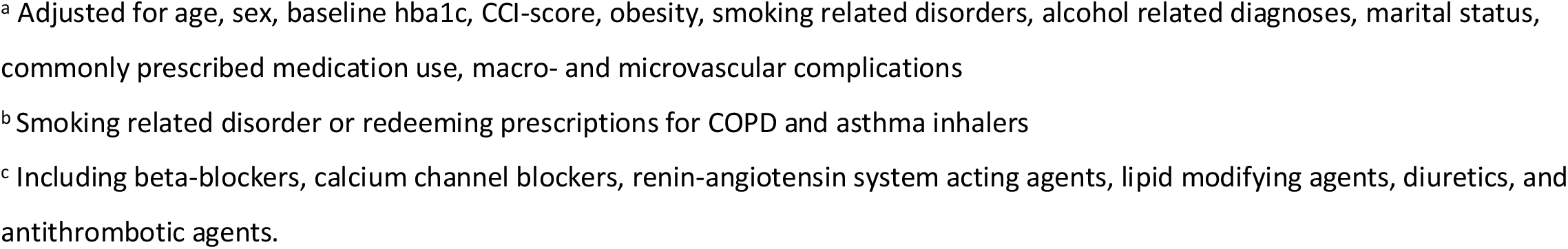
Association between baseline characteristics and initiation of psychopharmacological treatment/psychiatric hospital contact during follow up.

|  | Psychopharmacological treatment initiation |  | Psychiatric hospital contact |  |
| --- | --- | --- | --- | --- |
|  | Unadjusted HRR | Fully adjusted HRR<sup>a</sup> | Unadjusted HRR | Fully adjusted HRR<sup>a</sup> |
| Age |  |  |  |  |
| <=50 | 1.00 | 1.00 | 1.00 | 1.00 |
| 51-60 | 0.86 (0.78-0.95) | 0.84 (0.76-0.93) | 0.37 (0.28-0.50) | 0.39 (0.28-0.53) |
| 61-70 | 0.89 (0.81-0.98) | 0.82 (0.74-0.91) | 0.24 (0.17-0.32) | 0.24 (0.17-0.34) |
| 71-80 | 1.36 (1.24-1.49) | 1.15 (1.03-1.28) | 0.71 (0.55-0.91) | 0.73 (0.54-0.99) |
| >80 | 2.00 (1.81-2.21) | 1.46 (1.28-1.66) | 2.08 (1.66-2.61) | 1.98 (1.43-2.75) |
| Sex |  |  |  |  |
| Female | 1.00 | 1.00 | 1.00 | 1.00 |
| Male | 0.91 (0.86-0.97) | 0.95 (0.89-1.01) | 0.78 (0.65-0.92) | 0.85 (0.70-1.02) |
| Hba1c |  |  |  |  |
| 6.5-7% | 1.00 | 1.00 | 1.00 | 1.00 |
| 7.1-7.5% | 1.01 (0.92-1.11) | 1.05 (0.95-1.16) | 0.85 (0.63-1.13) | 0.87 (0.64-1.17) |
| 7.6-8.5% | 1.06 (0.96-1.17) | 1.15 (1.04-1.27) | 0.91 (0.67-1.24) | 0.91 (0.66-1.25) |
| 8.6-9.5% | 0.96 (0.84-1.09) | 1.12 (0.98-1.28) | 0.93 (0.63-1.37) | 0.95 (0.63-1.43) |
| 9.6-10.5% | 0.92 (0.80-1.06) | 1.09 (0.94-1.26) | 0.86 (0.56-1.31) | 0.83 (0.52-1.32) |
| >10.5% | 0.87 (0.79-0.96) | 1.00 (0.90-1.11) | 0.86 (0.65-1.14) | 0.73 (0.53-1.02) |
| Charlsons Comorbidity index |  |  |  |  |
| 0 | 1.00 | 1.00 | 1.00 | 1.00 |
| 1 | 1.40 (1.31-1.50) | 1.23 (1.14-1.33) | 1.44 (1.18-1.75) | 1.37 (1.08-1.72) |
| 2 | 2.20 (2.05-2.37) | 1.67 (1.51-1.84) | 1.79 (1.42-2.26) | 1.63 (1.19-2.21) |
| Macrovascular complications | 1.55 (1.45-1.65) | 1.16 (1.06-1.27) | 1.35 (1.11-1.64) | 1.04 (0.80-1.35) |
| Microvascular complications |  |  |  |  |
| Overall | 1.66 (1.53-1.80) | 1.10 (1.01-1.21) | 1.85 (1.46-2.34) | 1.00 (0.77-1.31) |
| Eye | 1.64 (1.50-1.79) |  | 1.92 (1.50-2.46) |  |
| Renal | 1.56 (1.31-1.88) |  | 1.38 (0.78-2.45) |  |
| Atrial fibrillation | 1.62 (1.49-1.77) | 1.15 (1.04-1.26) | 1.49 (1.15-1.95) | 0.93 (0.68-1.26) |
| Obesity | 0.90 (0.80-1.02) | 0.94 (0.82-1.07) | 1.09 (0.77-1.53) | 1.20 (0.83-1.72) |
| Alcohol related diagnoses | 1.74 (1.47-2.06) | 1.69 (1.40-2.03) | 2.34 (1.51-3.61) | 2.19 (1.30-3.69) |
| Smoking related disorder<sup>b</sup> | 1.49 (1.39-1.60) | 1.17 (1.08-1.27) | 0.89 (0.69-1.15) | 0.72 (0.55-0.95) |
| Marital status |  |  |  |  |
| Married/partner | 1.00 | 1.00 | 1.00 | 1.00 |
| Unmarried | 0.69 (0.61-0.78) | 0.71 (0.62-0.81) | 1.36 (1.00-1.79) | 1.17 (0.86-1.61) |
| Divorced/ended partnership | 1.29 (1.17-1.43) | 1.22 (1.09-1.35) | 1.64 (1.22-2.22) | 1.68 (1.24-2.27) |
| Widow | 1.97 (1.81-2.13) | 1.35 (1.23-1.49) | 2.98 (2.38-3.73) | 1.38 (1.06-1.80) |
| Commonly prescribed medication use<sup>c</sup> |  |  |  |  |
| None | 1.00 | 1.00 | 1.00 | 1.00 |
| One | 1.09 (0.99-1.19) | 0.93 (0.85-1.03) | 0.97 (0.74-1.26) | 0.84 (0.63-1.13) |
| Two | 1.10 (1.03-1.17) | 0.76 (0.70-0.82) | 0.93 (0.77-1.12) | 0.68 (0.53-0.86) |

<sup>a</sup> Adjusted for age, sex, baseline hba1c, CCI-score, obesity, smoking related disorders, alcohol related diagnoses, marital status, commonly prescribed medication use, macro- and microvascular complications
<sup>b</sup> Smoking related disorder or redeeming prescriptions for COPD and asthma inhalers
<sup>c</sup> Including beta-blockers, calcium channel blockers, renin-angiotensin system acting agents, lipid modifying agents, diuretics, and antithrombotic agents.

The same characteristics were associated with psychiatric hospital contact (see Table 4), with the following exceptions: Having a macrovascular complication at baseline was not associated with a materially increased risk of psychiatric hospital contact (HR=1.04, 95%CI=0.80-1.35), and only individuals above the age of 80 had a clear statistically significant increased risk of psychiatric hospital contact (HR=1.98, 95%CI=1.43-2.75, while individuals aged 51-60 versus <=50 years (HR=0.39, 95%CI=0.28-0.53), 61-70 versus <=50 years (HR=0.24, 95%CI=0.17-0.34) and 71-80 (HR=0.73, 95%CI=0.54-0.99) were less likely to have a psychiatric hospital contact during follow-up compared to individuals under the age of 50 (see Table 4). The proportional hazard assumption was met in these analyses.

### Sensitivity Analyses

The results of the sensitivity analyses in which having the “outcome” was defined as redemption of at least two prescriptions (rather than only one) for psychopharmacological drugs within the two years following onset of T2D/matched date, were analogue to those of the main analyses. Specifically, the risk of requiring psychopharmacological treatment associated with T2D was also substantially increased in these analyses, including those stratified on antidepressants, antipsychotics and anxiolytics (see Supplementary Table 2 and 3).

## Discussion

In this register-based cohort study, we found that 8.3% of individuals with newly developed T2D initiated psychopharmacological medication use within 2 years from their diagnosis compared to 4.6% among age and sex matched controls from the general population. Furthermore, in analyses using a propensity score matched design to minimize confounding, we found that individuals with newly developed T2D were at increased risk of initiating psychopharmacological treatment, compared to controls. Finally, we identified a number of risk factors, which were associated with elevated risk of requiring psychopharmacological treatment or psychiatric hospital contact in the two years following the onset of T2D, including older age, somatic comorbidity, and being divorced/widowed

### Incidence rates of psychopharmacological treatment and psychiatric hospital contact

The finding from our current study, namely that 5.6% and 2.9% of the individuals with newly developed T2D initiated antidepressant- and anxiolytic treatment, respectively, within two years, is somewhat contrasting to prior studies in the field (incidence of 7.10-10.4% for antidepressant treatment and 6.5% for anxiolytic treatment) (Mast et al., 2017, Perez et al., 2017, Cleal et al., 2018, Kivimäki et al., 2010). However, in contrast to most other studies, we excluded individuals having received psychopharmacological treatment or having had a psychiatric hospital contact within the five years leading up to the onset of T2D, in order to get better estimates of the true incidence of the need for psychopharmacological treatment and specialist psychiatric care (hospital contact) among individuals with newly developed T2D. When comparing the incidence of psychopharmacological treatment and psychiatric hospital contact in T2D to that of age and sex matched controls, the incidence rates were substantially higher among those with T2D. This is however not necessarily due to T2D per se, as this condition is associated with a range of characteristics that increase the risk of mental illness – including somatic comorbidities (Nowakowska et al., 2019, Adriaanse et al., 2016) and unhealthy lifestyle (Kolb and Martin, 2017, Schellenberg et al., 2013) – that are likely to confound the association between T2D and the outcomes studied here.

### The association between T2D and psychopharmacological treatment and psychiatric hospital contact

In order to minimize the degree of confounding in the estimation of the association between newly developed T2D and psychopharmacological treatment/psychiatric hospital contact, we conducted a propensity score matched analysis, which confirmed a 51% increased risk of psychopharmacological treatment in T2D. While we can by no means claim causality based on these findings, they are compatible with T2D increasing the risk for mental disorders. The potential mechanisms underlying such an effect may include, but are not limited to, the psychological distress associated with developing T2D (Chew et al., 2017, Aljuaid et al., 2018), a potential direct effect of glycemia on the brain (Giri et al., 2018, Kim et al., 2016), and downstream effects of T2D such as cardiovascular disease (Zheng et al., 2018, Gedebjerg et al., 2018). This study was however not designed to investigate the individual contributions of (and potential interaction between) these potential mechanisms.

As opposed to the findings for psychopharmacological treatment, the risk of psychiatric hospital contact was only increased by 14% for those with T2D in the propensity score matched analysis. A possible explanation for this difference – linking the apparently opposing findings – is that those in need of treatment for mental disorders receive relevant psychopharmacological treatment by their general practitioner (Musliner et al., 2019) and do therefore not go on to develop conditions so severe that treatment/assessment at psychiatric hospitals is required.

### Risk factors for psychopharmacological treatment and psychiatric hospital contact in T2D

In the analysis of which baseline characteristics that were associated with subsequent psychopharmacological treatment and psychiatric hospital contact among individuals with T2D, we found that older age, somatic comorbidity, and being divorced or widowed were risk factors for both psychopharmacological treatment and psychiatric hospital contact in individuals with newly developed T2D. These results are in agreement with risk factor studies from the field of depression (Schaakxs et al., 2017, Copeland et al., 2004, Scott et al., 2010) and suggest that clinicians should be particularly aware of development of mental disorder (and probably depression in particular) in individuals who are elderly, divorced/widowed and suffer from comorbid somatic illness at the onset of T2D.

Interestingly, individuals with T2D receiving other commonly prescribed medications at baseline were less likely to initiate psychopharmacological treatment. A potential explanation for this finding may be that these individuals are generally compliant to pharmacological treatment and may therefore also be adherent to potential glucose lowering medications, which may – in turn – result in well-regulated T2D and reduced risk of developing mental disorder requiring pharmacological/psychiatric hospital treatment.

## Limitations

Our findings should be interpreted in the light of the following limitations. First, due to the known comorbidity between T2D and mental illness, the identification of T2D (a HbA1c level >6.5%) itself might lead to the identification of mental illness and thereby psychopharmacological treatment initiation/psychiatric hospital contact. If such an ascertainment bias has affected this study, it would result in an overestimation of the strength of the association between T2D and the requirement for psychopharmacological treatment/psychiatric hospital contact. Second, although we primarily consider psychopharmacological treatment as a proxy for development of mental disorder, a proportion of the individuals with T2D will likely have initiated treatment with an antidepressant (Serotonin–norepinephrine reuptake inhibitors and tricyclic antidepressants in particular) due to neuropathic pain developed as a complication to T2D (Gilron et al., 2015). However, as we only included individuals with newly developed T2D, only a relatively small proportion will have developed neuropathic pain requiring pharmacological treatment after two years. Also, the positive association between T2D and psychopharmacological treatment was observed across all investigated groups of drugs in this category – and not only those that are typically used in the management of neuropathic pain. Third, the data from this study stems from the Danish national healthcare system, which provides tax-funded healthcare for all citizens. Hence, the results may not necessarily translate to other countries – especially those with healthcare systems based on other models. Therefore, the results should ideally be replicated in other settings to confirm generalizability.

## Conclusion

Individuals with newly developed T2D are at elevated risk of requiring psychopharmacological treatment compared to propensity score matched controls without T2D. Risk factors for psychopharmacological treatment/psychiatric hospital contact following development of T2D are consistent with those observed for general populations and include older age, somatic comorbidity and being divorced or widowed.

## Data Availability

No data available.

## Acknowledgements

None

## Author contributions

The study was designed in collaboration between all authors. CR conducted the analyses. The results were interpreted by all authors. The manuscript was drafted by CR and SDØ and revised for important intellectual content by NS and RWT. The final version was approved by all authors prior to submission.

## Supplementary information

**Supplementary table 1:**
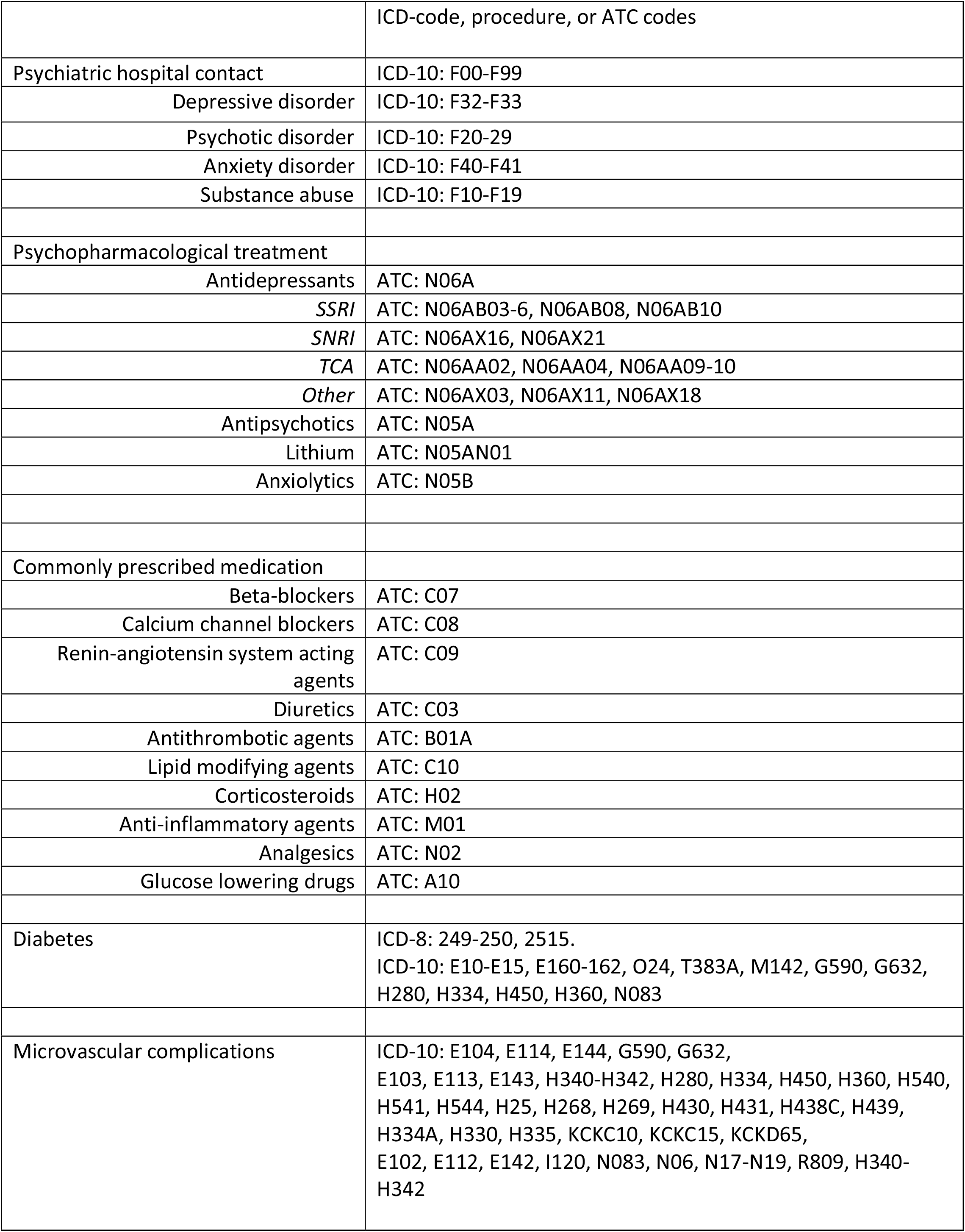
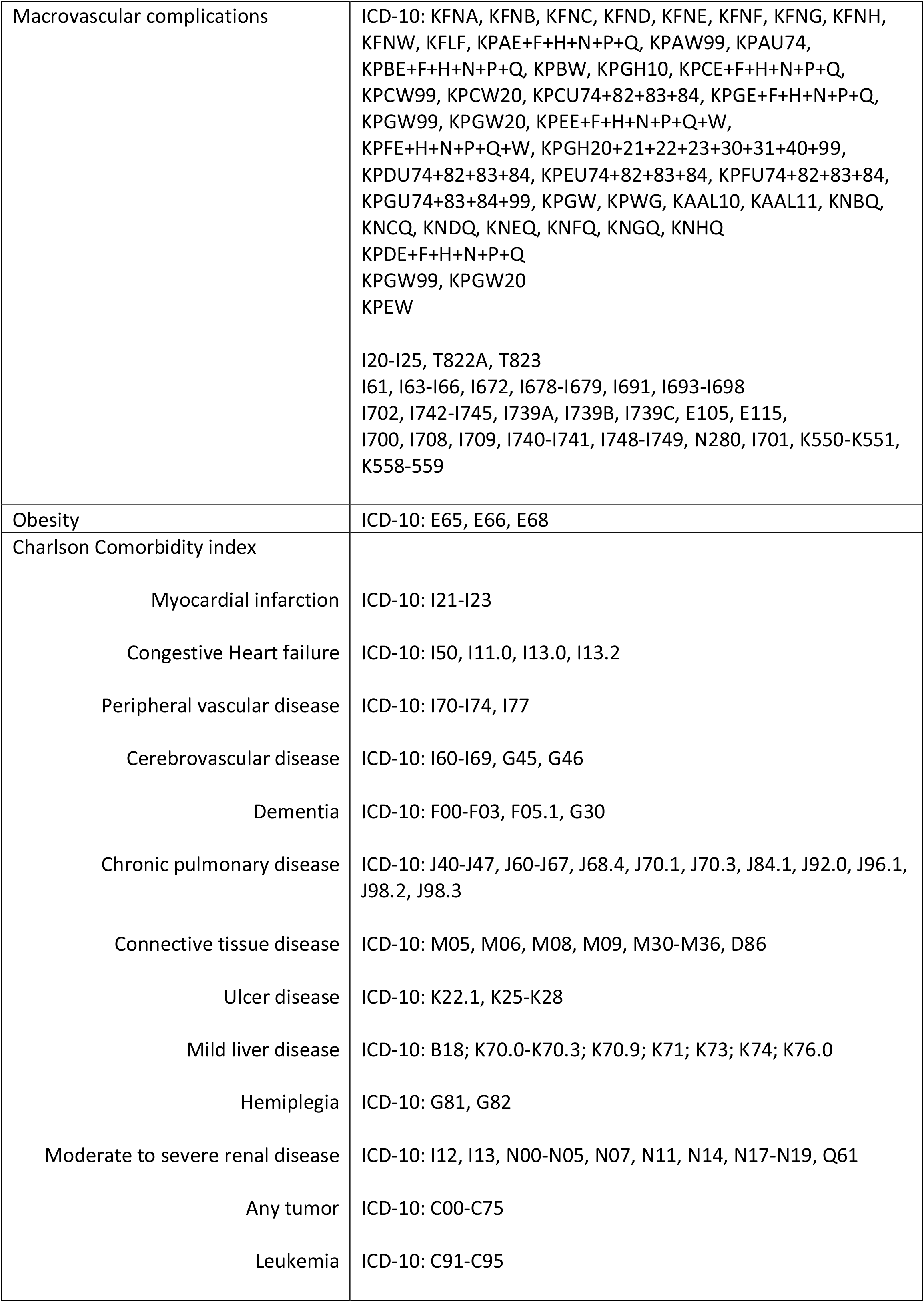
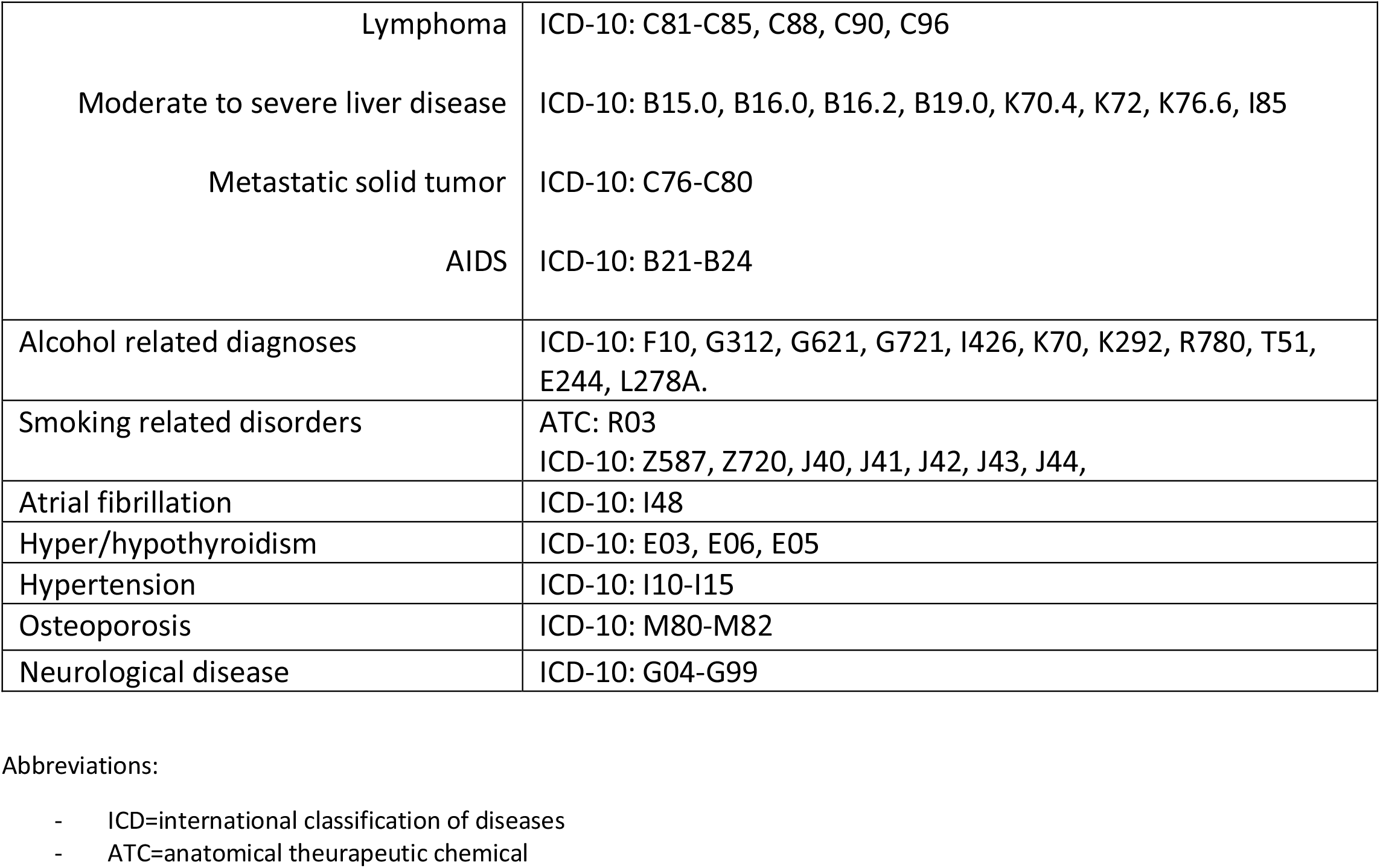
Definition of variables based on ATC, ICD-8, ICD-10, and procedure codes.

**Supplementary table 2:** Use of psychopharmacological treatment within the first two years after incident T2D or index date

|  | Individuals with T2D<br>N=56,640 (%) |  | Individuals without T2D (controls)<br>N=315,694 (%) |  |
| --- | --- | --- | --- | --- |
|  | Number of<br>individuals (%) | Incidence rate pr<br>1000 years follow-up,<br>(95% CI) | Number of<br>individuals (%) | Incidence rate pr 1000<br>years follow-up,<br>(95% CI) |
| **Any psychopharmacological agent** | 2,830 (5.0%) | 26.38 (25.43-27.37) | 7,835 (2.5%) | 12.65 (12.37-12.93) |
| **Antidepressants** |  |  |  |  |
| Any antidepressant | 2,155 (3.8%) | 20.09 (19.26-20.96) | 5,553 (1.8%) | 8.96 (8.73-9.20) |
| SSRI | 1,222 (2.2%) | 11.39 (10.77-12.05) | 3,091 (1.0%) | 4.99 (4.82-5.17) |
| SNRI | 174 (0.3%) | 1.62 (1.40-1.88) | 302 (0.1%) | 0.49 (0.44-0.55) |
| TCA | 351 (0.6%) | 3.27 (2.95-3.63) | 729 (0.2%) | 1.18 (1.09-1.27) |
| Other | 459 (0.8%) | 4.28 (3.90-4.69) | 1,389 (0.4%) | 2.13 (2.13-2.36) |
| **Antipsychotics** |  |  |  |  |
| Any antipsychotic | 381 (0.7%) | 3.55 (3.21-3.93) | 902 (0.3%) | 1.46 (1.36-1.55) |
| **Anxiolytics** |  |  |  |  |
| Any anxiolytic | 615 (1.1%) | 5.73 (5.30-6.20) | 2,236 (0.7%) | 3.61 (3.46-3.76) |

Abbreviations: T2D=type 2 diabetes, SSRI=selective serotonin reuptake inhibitor, SNRI=serotonin–norepinephrine reuptake inhibitor, TCA=tricyclic antidepressant

**Supplementary table 3:** Use of psychopharmacological treatment within the first two years after incident T2D or index date, using propensity score matching

|  | Propensity score model |  |  |
| --- | --- | --- | --- |
|  | Individuals with T2D<br>(n=44,742) | Propensity score matched<br>controls without T2D<br>(n=44,742) | Hazard ratio (HR) |
| **Any psychopharmacological agent** | 2,105 (4.7%) | 1,335 (3.0%) | 1.63 (1.52-1.74) |
| **Antidepressants** |  |  |  |
| Any antidepressant | 1,598 (3.6%) | 992 (2.2%) | 1.66 (1.54-1.80) |
| SSRI | 891 (2.0%) | 504 (1.1%) | 1.82 (1.63-2.03) |
| SNRI | 127 (0.3%) | 63 (0.1%) | 2.07 (1.53-2.80) |
| TCA | 269 (0.6%) | 148 (0.3%) | 1.87 (1.53-2.28) |
| Other | 350 (0.8%) | 268 (0.6%) | 1.34 (1.14-1.57) |
| **Antipsychotics** |  |  |  |
| Any antipsychotic | 273 (0.6%) | 144 (0.3%) | 1.94 (1.59-2.38) |
| **Anxiolytics** |  |  |  |
| Any anxiolytic | 472 (1.1%) | 340 (0.8%) | 1.42 (1.24-1.64) |

Abbreviations: T2D=type 2 diabetes, SSRI=selective serotonin reuptake inhibitor, SNRI=serotonin–norepinephrine reuptake inhibitor, TCA=tricyclic antidepressant

